# The association between the value of a statistical life and frailty in Burkina Faso

**DOI:** 10.1101/2024.02.10.24302634

**Authors:** Leila Freidoony, Dina Goodman-Palmer, Fred Barker, Mamadou Bountogo, Pascal Geldsetzer, Guy Harling, Lisa Hirschhorn, Jennifer Manne-Goehler, Mark Siedner, Stefan Trautmann, Yilong Xu, Miles Witham, Justine Davies

## Abstract

**Background:** To ensure resources invested into services are commensurate with benefit, economists utilise various methods to assess value of life. Understanding the performance of these methods in older populations is crucial, particularly in low-and-middle-income countries (LMICs), where the majority of older people will live by 2030. Value of Statistical Life Years (VSLY) is widely used in cost-benefit analyses but rarely been in LMICs or in older people.

**Objective:** This study aimed to investigate the hypothesis that frailty would be associated with a lower VSLY in participants in rural Burkina Faso, when controlling for factors found in other studies likely to affect VSLY, such as socio-demographics, multimorbidity, quality of life, and disability.

**Methods:** The study included 3,988 adults aged 40 years and older from a population-representative household survey done in Nouna, Burkina Faso. Data were collected on sociodemographic characteristics, chronic medical conditions, quality of life, disability, physical performance, and VSLY. Frailty status was derived using Fried’s frailty phenotype. Bivariate analyses investigated the association between quintiles of VSLY and frailty. To explore modification of associations by other variables, we built sequential binary logistic regression models comparing each quintile of VSLY with the first (lowest) quintile. Models included frailty category, age, sex, marital status, educational attainment, and wealth. We sequentially added quality of life, multimorbidity, and disability.

**Results:** Of 2,761 survey participants included in this analysis, 51.4% were female. Average age was 54.5 years (with 70.0% aged 40-59 years), 24.8% of respondents reported being alone, and 84.5% had not completed education. In bivariate analyses, we found a significant negative association between higher VSLY and frailty, increasing age, disability, and quality of life. Conversely, being male, married, and educated were positively associated with higher VSLY. The negative association between VSLY and frailty remained significant after adjusting for age, gender, education, wealth, quality of life, disability, and multimorbidity (odds of being frail for VSLY quintile 5 vs quintile 1 was 0.48, 95% CI 0.37-0.64 for the fully adjusted model). Furthermore, effect of age, education, and wealth on VSLY became non-significant once frailty was included in the model.

**Conclusion:** There is a strong relationship between the value that older people place on their lives and their frailty status. Frailty status is important to consider when assessing VSLY, especially in LMICs in which there is a rapidly growing older population.

**Key Points for Decision Makers:**

- This study explores the relationship between frailty and the Value of Statistical Life Years (VSLY) in older adults in rural Burkina Faso, representing the first such investigation in any setting.
- The research reveals a strong and significant association between frailty and lower VSLY, even after adjusting for variables like age, multimorbidity, and quality of life.
- These findings emphasize the importance of considering frailty status in the application of VSLY in cost-benefit analyses, particularly for interventions targeting older adults in Low- and Middle-Income Countries.

## 1. Introduction

To ensure that resources invested into services are commensurate with their benefit, economists have utilised various methods to assess the value of life. As populations age, it is important that the performance of these methods is understood in older people. This is especially the case for older populations living in low- and middle-income countries (LMICs), given that by 2030, the majority of the world’s older people are expected to be living in LMICs [1]. The Value of Statistical Life Years (VSLY) approach has consistently calculated human life as being far more valuable than alternative methods of cost-benefit analyses (CBAs) such as ‘value of lost earnings’ or the World Health Organization’s (WHO) cost-effectiveness threshold of multiplying national GDP per capita by three [2, 3]. The economic theory underlying the higher VSL measurements raises the possibility that previous methods have undervalued human life, hence its interest to health economists.

VSLY is a commonly used tool specific to CBAs where risk of death is considered as part of evaluation. It is guided by the real-world or theoretical monetary value that individuals place on accepting or reducing a small risk of death, either over their lifetime or annually [3]. The generated values essentially behave as a monetary threshold beyond which additional annual expenditure is considered excessive for the marginal reduced risk of death it provides. Whilst discussion continues on the benefits of using it, the VSLY approach continues to be viewed as beneficial, at least in high income countries [4, 5]. The few studies which have used the approach in sub-Saharan Africa or in LMICs in general have also considered it to be worthwhile [6]. But, given the growing ageing population in LMICs, understanding the impact of ageing on VSLY is needed [7], particularly if this is to be considered an applicable method for LMICs.

There is reasonable evidence from high-income countries showing the relationship between VSL values and increasing age [8]. However, there is a paucity of research exploring whether these findings are consistent in lower-income countries where life expectancy is typically lower and the role of older people in their communities is often more prominent [9, 10]. There is also little evidence on the effect of frailty on VSLY as people age. Frailty is a condition describing an individual’s vulnerability to greatly impaired health status following relatively minor stressor events [11]. Moreover, frailty occurs more frequently as people age and is associated with limiting independence and reduced quality of life; it is prevalent in older populations in LMICs [12–14].

Given the increase in older populations in LMICs, the high prevalence of frailty and its adverse consequences in these populations, and that VSLY is an attractive method to understand the value of life in low resourced settings, it is important to understand the relationship between frailty and VSLY [9, 15, 16]. Our hypothesis was that frailty would be associated with a lower VSLY in participants in rural Burkina Faso. This study aims to test our hypothesis, when controlling for factors found in other studies likely to affect VSLY [17–19], such as, socio-demographics, multimorbidity, quality of life, and disability.

## 2. Methods

### 2.1. Study setting/Population description

We enrolled older adults (aged 40 and over) living in the Nouna Health and Demographic Surveillance System (HDSS) area in north-western Burkina Faso. Burkina Faso is one of the poorest countries in the World, with a Gross Domestic Product of $893.08 (2021), ranking 184^th^ on the Human Development Index.

The Centre de Recherche en Santé de Nouna (CRSN) led household surveys within the Nouna HDSS, which consists of the town of Nouna and 59 surrounding villages, with a total population of 107,000 residents from multiple religious backgrounds. There are two main languages spoken in Nouna— Dioula and Moore—whereas French is the national language. The main economic activity is subsistence farming. Details of study design is described elsewhere [15, 20, 21].

### 2.2. Sample

Data for this study were collected during the baseline wave of the CRSN Heidelberg Aging Study, which was conducted in 2018, using a household survey from a population-representative sample of adults ≥40 years living in the Nouna HDSS area. To obtain a sample of 3000 older adults, we used a two-part multistage random sample of 4000 individuals from the 2015 Nouna HDSS census, allowing for 25% loss due to mortality, mobility, or non-response. In the six villages with fewer than 50 adults aged over 40, all adults were selected to take part. In all other villages, a random sample of households with at least one person over 40 was drawn, and then within each selected household one age-eligible adult was randomly selected to complete the survey. Details of study design can be found elsewhere [15, 20, 21].

### 2.3. Data collection

Trained data collectors administered the survey to participants in the local language. The household survey questionnaire has been described in full in other publications [22–24]. It included questions on sociodemographic characteristics, health and medical conditions, physical performance, and VSLY which were used in this study.

#### 2.3.1. Sociodemographic characteristics

We captured sociodemographic characteristics, including age in years; gender (*male or female*); marital status (*never married*, *widowed*, *married but separated/divorced*, *currently married, or cohabiting*); educational attainment (*no formal schooling, less than primary, primary complete, some secondary, secondary complete, high school complete, or college/university)*; and a suite of 37 questions on household assets and dwelling characteristics.

#### 2.3.2. Chronic conditions

Self-reported chronic conditions collected in the study were communicable (HIV) or non-communicable health conditions (cancer, chronic respiratory disease, stroke, heart disease, hypertension, diabetes, and hypercholesterolaemia). Cognitive impairment was assessed using the eight-question community screening interview for dementia (CSI-D), anxiety using the Generalised Anxiety Disorder question (GAD-2) score, and depression using the Patient Health Questionnaire (PHQ-9) [25]. Additionally, questions from the Centre for Epidemiologic Studies Depression Scale (CES-D) were used to ask questions related to levels of exhaustion; ‘Everything I did in the last week was an effort’ or ‘I could not get going’ [26]. Quality of life was asked using the EuroHIS 8-item version of the WHO Quality of Life (WHOQOL) questionnaire [27].(14). Disability was measured using the 12-item WHO Disability Assessment Schedule, Version 2 (WHODAS V.2.0) disability score. This score measures impairments in function, activity, and participation (mobility, self-care, cognition, interaction with others, life activities, and social participation), with higher scores indicating higher levels of disability [28].

#### 2.3.3. Physical measurements

We measured weight and height using generic scales and measurement bands against a vertical and horizontal smooth surface, respectively. Blood pressure of participants was measured in a seated position from the left arm after 15 min rest; three measures were taken using Omron Series 7 portable blood pressure machines (Omron Healthcare, Kyoto, Japan). The mean of the second and third measurements, taken at least 5 min apart, were used in the analysis.

Walk speed was measured using a digital stopwatch and recorded to the nearest tenth of a second with the participant walking 4m course marked out on level ground, twice. The fastest speed in metres per second (m/s), adjusted for height, was used. Hand grip strength was measured using a Jamar Hydraulic Hand Dynamometer. We took measurements while the participant seated, the arm at 90° elbow flexion and the shoulder and wrist in the neutral position. Two attempts were recorded from each hand; the maximum was used in this analysis.

#### 2.3.4. Blood tests

Capillary glucose levels were analysed using SD Biosure Code free point-of-care testing strips, and cholesterol using the Jactron Pictus 400 machine.

#### 2.3.5. VSLY

VSLY was assessed using four related methods; willingness to pay or willingness to accept a hypothetical mortality risk reduction or increase, with each using two different ranges of payments – large or small (15, 16). We elicited the annual willingness-to-pay (WTP) (or willingness-to-accept, WTA) for a 2% reduction (or increase) in risk of death from a 5% to a 3% level, or from a 3% to a 5% level. The 5% risk represents the typical mortality rate in Burkina Faso, as determined by life tables for 2015. We calculated the VSLY directly from the questionnaire. The observed WTP/WTA was the value of a 2% absolute risk reduction/increase; so, the VSLY was obtained by multiplying this elicited WTP/WTA measurement by 50 to get to the annual value of a 100% reduction/increase in risk of death [6].

After each participant was randomised to either WTP or WTA, we factorially randomized them to receive either a small or large range of payment amounts as response options, both starting at zero Communauté Financière Africaine franc (West African CFA franc). The small range of payments had an upper limit of 400,000 CFA (about US$ 1916) and the large range, an upper limit of 2 million CFA (about US$ 9580). For comparison, gross domestic product per capita at purchasing power parity in Burkina Faso was US$2,044 in 2017, when the experiment was designed. See Supplementary Information 1 for details.

### 2.4. Definition of variables

#### 2.4.1. VSLY

We derived the outcome variable VSLY as quintiles for each of the four methodologies (WTP-small, WTP-large, WTA-small, and WTA-large), and then quintiles were harmonised across all participants. The continuous values from each of the four VSLY methods were also used.

#### 2.4.2. Frailty

Frailty was derived based on Fried’s frailty score and constructed using five domains (weight loss, activity levels, exhaustion, grip strength, and walk speed) that were previously validated in LMICs [15, 16, 20, 29]. Variables and thresholds to create the frailty categories are summarised in Supplementary Information 2. Individuals were categorised as non-frail/robust (0 points), pre-frail (1– 2 points) or frail (3–5 points) [30]. Frailty was then dichotomized as frail or pre-frail compared to non-frail [16].

#### 2.4.3. Other covariates

Marital status was categorised as either ‘single’ (*never married*/*widowed*/*married but separated/divorced*) or ‘married’ (*currently married/cohabiting*). Education was categorised as ‘no education’ or ‘any education’. Wealth quintiles were derived from the questions on household assets and dwelling characteristics using the Principal Component Analysis method of Filmer and Pritchet [31].

Multimorbidity was defined as having two or more chronic conditions (HIV, cancer, chronic respiratory disease, stroke, heart disease, hypertension, diabetes, hypercholesterolaemia, cognitive impairment, depression, or anxiety). As is common in epidemiology studies, hypertension was defined as either self-reported previous diagnosis or currently being on treatment, or as a measured systolic blood pressure ≥140 mm/Hg, or a diastolic blood pressure ≥90 mm/Hg; diabetes was defined as either self-reported diagnosis, being on treatment, or by blood test (a non-fasting point of care capillary glucose level >200 mg/dL, HbA1c>6.5% or fasting glucose >126 mg/dL); hypercholesterolaemia was defined as self-reported diagnosis; being on treatment; or by blood test (a total plasma cholesterol >200 mg/dL, low density lipoprotein >160 mg/dL).

Quality of life was derived by summing responses to WHOQOL before normalising to a 0–100 scale, with 100 denoting the best quality of life. Disability measures were also summed and then normalised to a 0-100 scale, where 100 represents the worst disability [27, 28, 32]. Participants were defined as having symptoms of anxiety if their GAD-2 score was ≥3, those with PHQ-9 of ≥10 were categorised as having depressive symptoms, and those with a CSI-D score of less than 7 were categorised as having possible/probable cognitive impairment.

### 2.5. Statistical analysis

Continuous variables were described as mean and standard deviation (SD) or median and interquartile range (IQR) [25^th^/75^th^ centiles] if not normally distributed. Categorical variables were described as count (n) and proportion (%).

Frailty was the primary independent variable. Other independent variables we hypothesised to be associated with VSLY were included in the adjusted analyses: age, gender, marital status, education level, wealth, quality of life, multimorbidity, and disability. VSLY was the primary outcome.

We conducted four sets of analyses to elucidate the relationship between VSLY and frailty. First, we assessed the association between VSLY as a continuous variable and frailty category in each of the four individual VSLY methodological arms using Mann-Whitney U tests.

Second, bivariate associations between VSLY quintiles and frailty categories and each individual variable (age, sex, marital status, education status, wealth, quality of life, multimorbidity, and disability) were tested using Pearson’s chi-squared test, independent samples t-test, or one-way ANOVA, depending on the data type and distribution.

Third, to test whether variables were independently associated with VSLY, and to explore whether any associations between frailty and VSLY quintiles were modified by other variables, after excluding collinearity between variables, we built sequential binary logistic regression models comparing each quintile of VSLY with the first quintile. In each model, we included frailty, socio-demographic and economic characteristics of age, sex, marital and education status, and wealth; to these variables, we sequentially added quality of life, multimorbidity, and disability.

Fourth, we conducted a multinomial regression as a sensitivity analyses, with all VSLY quintiles included, fully adjusted for frailty and other covariates.

Results of analyses were presented using tables and forest plots displaying Odds Ratios (OR) and 95% confidence intervals (CI). We conducted a complete case analysis. All analyses were done using SPSS V.28 (IBM Corp. Released 2021. IBM SPSS Statistics for Windows, Version 28.0. Armonk, NY: IBM Corp).

## 3. Results

We identified 3,998 individuals aged >40 and living in the HDSS site; of those, 3,027 (75.7%) were located and agreed to participate and completed the baseline interview. Of those, 2761 individuals were included in this analysis after excluding participants with missing variables (Figure 1). Overall, 696 (25.2%), 679 (24.6%), 690 (25.0%), and 696 (25.2%) participants were available for the WTP-small, WTP-large, WTA-small, and WTA-large, respectively (Figure 1). Participants’ characteristics for variables included in the analyses are presented in Table 1 and Table 2.

**Figure 1.**
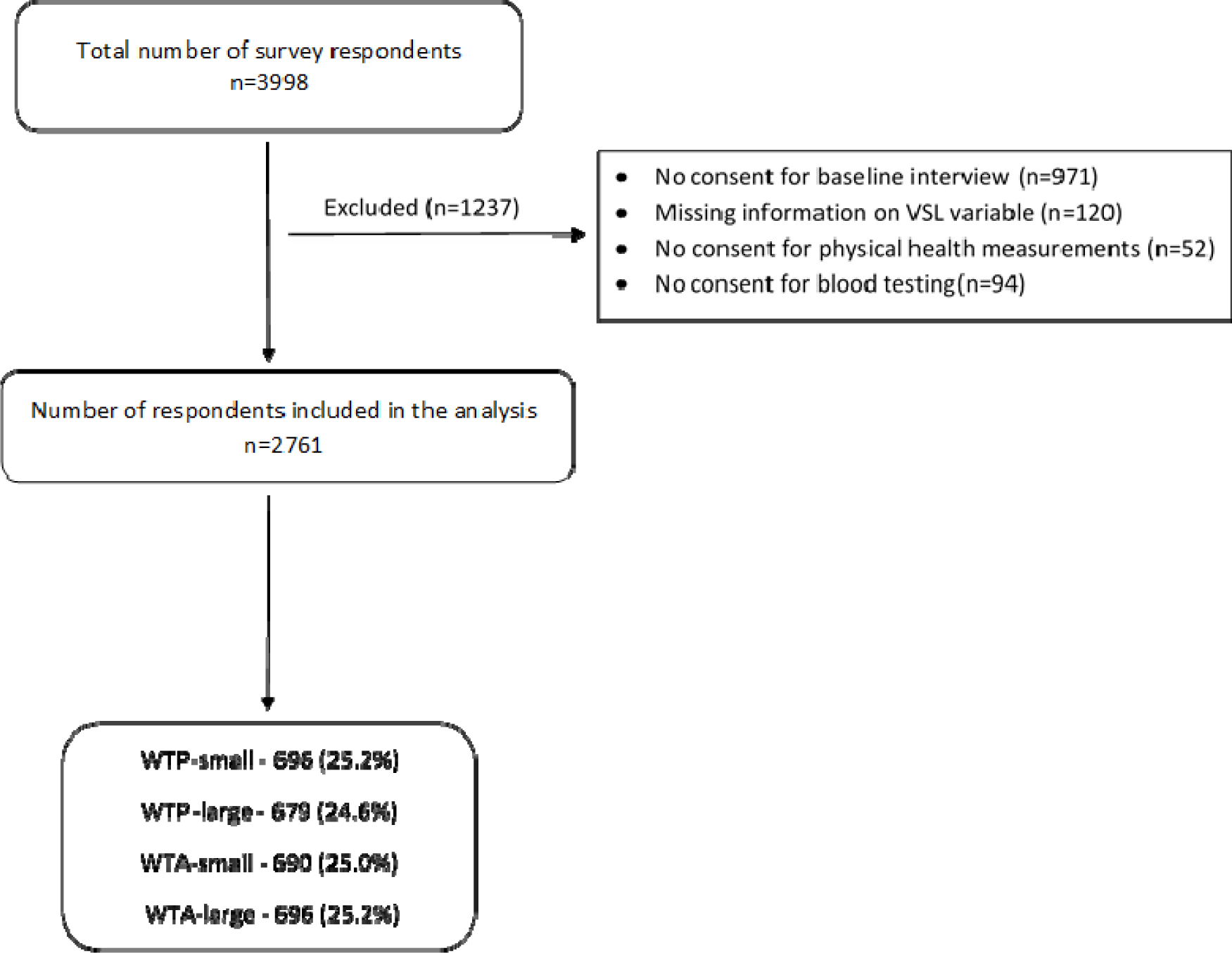
CONSORT flow diagram of participants included in final analyses. WTA, what to accept; WTP, what to pay.

**Table 1.** Baseline characteristics (n=2761)

| Parameter | Group | n (%), unless otherwise stated |
| --- | --- | --- |
| Gender (*n (%)*) | Male | 1341 (48.6) |
|  | Female | 1420 (51.4) |
| Age* (*Mean (SD)*) | - | 54.5 (11.0) |
| Age category (*n (%)*) | 40-49 | 1128 (40.9) |
|  | 50-59 | 802 (29.1) |
|  | 60-69 | 518 (18.8) |
|  | 70-79 | 249 (9.0) |
|  | 80+ | 62 (2.2) |
| Marital status (*n (%)*) | Widowed/divorced/single | 684 (24.8) |
|  | Married/cohabiting | 2077 (75.2) |
| Education level (*n (%)*) | No completed education | 2334 (84.5) |
|  | Some education | 427 (15.5) |
| Wealth quintile (*n (%)*) | 1 | 542 (19.6) |
|  | 2 | 548 (19.8) |
|  | 3 | 551 (20.0) |
|  | 4 | 569 (20.6) |
|  | 5 (most wealthy) | 551 (20.0) |
| Frailty (*n (%)*) | Non-frail | 1172 (42.4) |
|  | Pre-frail | 1227 (44.4) |
|  | Frail | 362 (13.1) |
| Multimorbidity (*n (%)*) | No | 2141 (77.5) |
|  | Yes | 620 (22.5) |
| Quality of life score (*Mean (SD)*) | - | 55.6 (14.3) |
| Disability score (*Mean (SD)*) | - | 15.0 (17.4) |
| VSLY arms (West African CFA franc)<br>*Median (25<sup>th</sup>-75<sup>th</sup> percentile)* | WTP-small | 5987.74<br>(5000.0-100000.0) |
|  | WTP-large | 11975.47<br>(10000.0-300000.0) |
|  | WTA-small | 16765.66<br>(20000.0-200000.0) |
|  | WTA-large | 59877.37<br>(50000.0-1000000.0) |
| Combined VSLY quintile (*n (%)*) | 1 | 563 (20.4) |
|  | 2 | 556 (20.1) |
|  | 3 | 546 (19.8) |
|  | 4 | 544 (19.7) |
|  | 5 (highest value) | 552 (20.0) |

\* *n*=2759 (2 missing values). VSLY, Value of Statistical Life Years

**Table 2.** Baseline characteristics by VSLY category, variables are presented as n (%), unless otherwise stated.

| Parameter |  | WTP small | WTP large | WTA small | WTA large |
| --- | --- | --- | --- | --- | --- |
| *N (% of total participants)* |  |  |  |  |  |
| Gender (*n (%)*) | Male | 329 (11.9) | 336 (12.2) | 336 (12.2) | 340 (12.3) |
|  | Female | 367 (13.3) | 343 (12.4) | 354 (12.8) | 356 (12.9) |
| Age* (*Mean (SD)*) | - | 54.8 (11.1) | 54.2 (11.2) | 54.7 (10.9) | 54.2 (10.8) |
| Age category (*n (%)*) | 40-49 | 276 (10) | 299 (10.8) | 259 (9.4) | 294 (10.6) |
|  | 50-59 | 203 (7.3) | 172 (6.2) | 227 (8.2) | 200 (7.2) |
|  | 60-69 | 136 (4.9) | 130 (4.7) | 128 (4.6) | 124 (4.5) |
|  | 70-79 | 61 (2.2) | 64 (2.3) | 57 (2.1) | 67 (2.4) |
|  | 80+ | 20 (0.7) | 14 (0.5) | 18 (0.6) | 10 (0.4) |
| Marital status (*n (%)*) | Widowed/divorced /single | 182 (6.6) | 165 (6.0) | 172 (6.2) | 165 (6.0) |
|  | Married/cohabitating | 514 (18.6) | 514 (18.6) | 518 (18.8) | 531 (19.2) |
| Education level (*n (%)*) | No completed education | 582 (21.1) | 581 (21.0) | 578 (20.9) | 593 (21.5) |
|  | Some education | 114 (4.1) | 98 (3.5) | 112 (4.0) | 103 (3.7) |
| Wealth quintile (*n (%)*) | 1 | 144 (5.2) | 138 (5.0) | 136 (4.9) | 124 (4.5) |
|  | 2 | 140 (5.1) | 138 (5.0) | 132 (4.8) | 138 (5.0) |
|  | 3 | 132 (4.8) | 141 (5.1) | 141 (5.1) | 137 (5.0) |
|  | 4 | 140 (5.1) | 144 (5.2) | 143 (5.2) | 142 (5.1) |
|  | 5 (most wealthy) | 140 (5.1) | 118 (4.3) | 138 (5.0) | 155 (5.6) |
| Frailty (*n (%)*) | Non-frail/Robust | 285 (10.3) | 270 (9.8) | 321 (11.6) | 296 (10.7) |
|  | Pre-frail | 310 (11.2) | 322 (11.7) | 283 (10.2) | 312 (11.3) |
|  | Frail | 101 (3.6) | 87 (3.1) | 86 (3.1) | 88 (3.2) |
| Multimorbidity (*n (%)*) | No | 513 (18.6) | 542 (19.6) | 541 (19.6) | 545 (19.7) |
|  | Yes | 183 (6.6) | 137 (5.0) | 149 (5.4) | 151 (5.5) |
| Quality of life score (*Mean (SD)*) | - | 55.5 (14.0) | 55.3 (14.4) | 55.6 (14.6) | 56.0 (14.2) |
| Disability score (*Mean (SD)*) | - | 16.1 (18.1) | 14.0 (17.4) | 15.6 (17.6) | 14.2 (16.4) |
| VSL arms (West African CFA franc) (*Median (IQR)*) | - | 5987.7 (22753.4) | 11975.5 (69457.7) | 16765.7 (95803.8) | 59877.4 (227534.0) |
| Combined VSLY quintile (*n (%)*) | 1 | 141 | 136 | 140 | 146 |
|  | 2 | 142 | 139 | 137 | 138 |
|  | 3 | 137 | 134 | 136 | 139 |
|  | 4 | 137 | 133 | 140 | 134 |
|  | 5 (highest value) | 139 | 137 | 137 | 139 |

VSLY, Value of Statistical Life Years; WTA, what to accept; WTP, what to pay

Significant, negative associations were seen between VSLY and frailty when considering each VSLY arm separately as a continuous variable (See Table 3), with the median willingness to pay value being around double for people in the non-frail compared with the frail or pre frail group.

**Table 3.** Median (IQR) and 25th/75th centiles of VSLY (West African CFA franc) as a continuous variable, for each of the 4 VSLY methodological arms - by frailty status.

| VSLY arms | Non-frail<br>Median (IQR)<br>[25<sup>th</sup> / 75<sup>th</sup> centiles] | Pre-frail/Frail<br>Median (IQR)<br>(25<sup>th</sup> / 75<sup>th</sup> centiles) | *p*-value* |
| --- | --- | --- | --- |
| WTP-small (n=696) | 8382.8 (22753.4)<br>[5000.00 / 100000.00] | 4790.2 (17963.2)<br>[5000.00 / 80000.00] | 0.006 |
| WTP-large (n=679) | 23950.9 (117359.7)<br>[10000.00 / 500000.00] | 11975.5 (58081.1)<br>[75000.00 / 250000.00] | 0.049 |
| WTA-small<br>(n=690) | 23950.9 (38321.5)<br>[40000.00 / 200000.00] | 11975.5 (32333.8)<br>[15000.00 / 150000.00] | <0.001 |
| WTA-large (n=696) | 107779.3 (227534.0)<br>[50000.00 / 1000000.00] | 47901.9 (161668.9)<br>[25000.00 / 700000.00] | <0.001 |

\* *p*-value comparing medians for each VSLY arm by frailty status using Mann-Whitney U test

In bivariate analyses, there were significant negative associations between the combined VSLY quintiles for the whole sample and frailty; those pre-frail/frail participants were less likely than non-frail ones to report higher VSLY (See Figure 2). Being male, married, and educated were significantly positively associated with increasing VSLY quintiles. Increasing age, disability, and quality of life, were negatively associated with increasing VSLY quintile. We saw no association between wealth and VSLY (Table 4).

**Figure 2.**
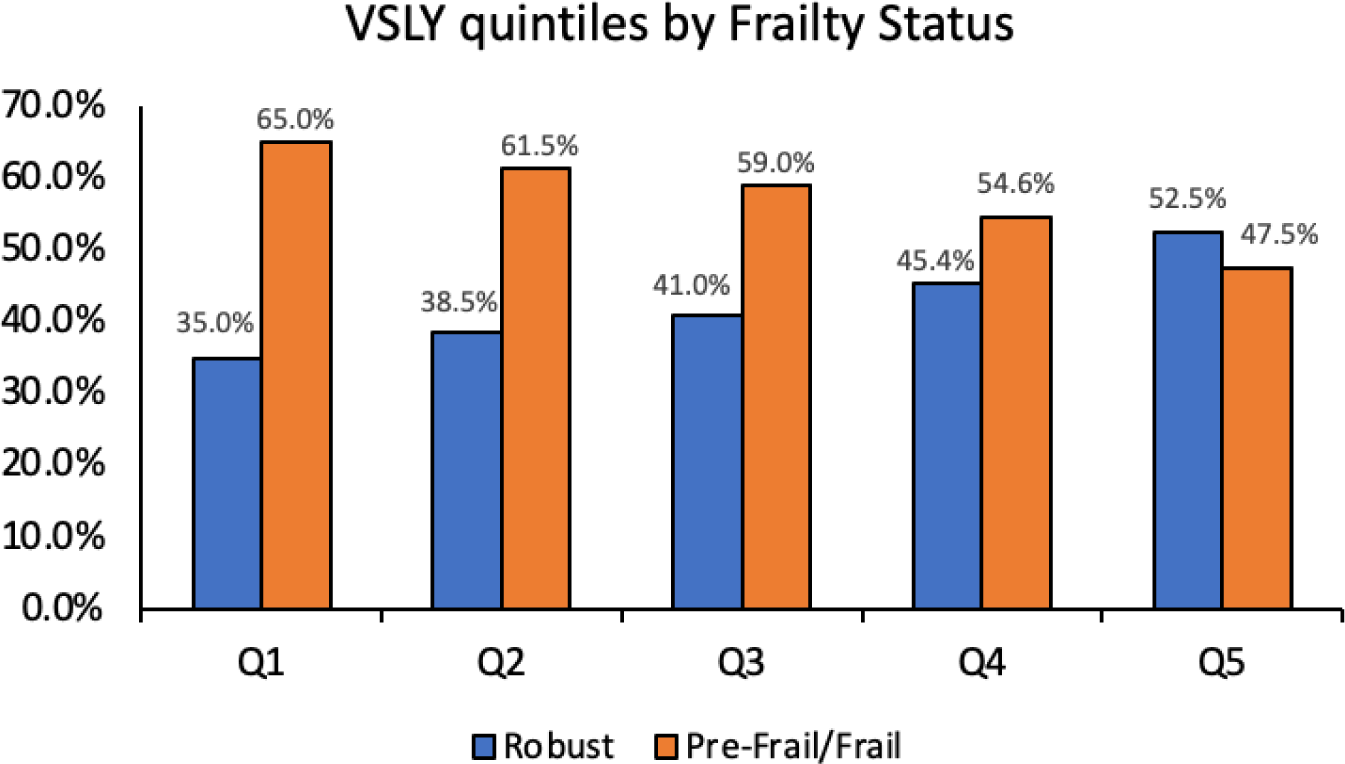
Association of frailty status and VSLY quintiles. Q, Quintile.

**Table 4.**
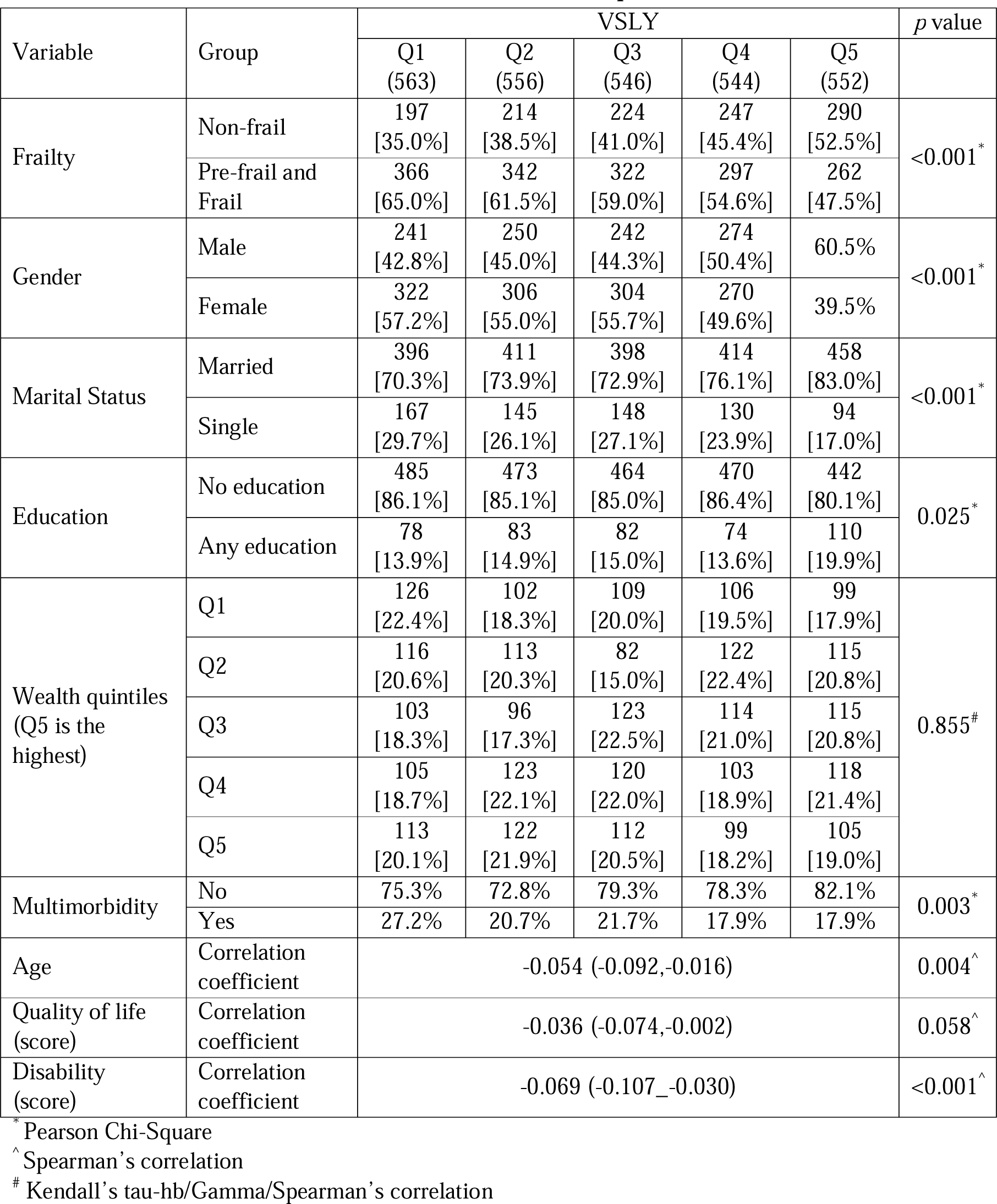
Bivariate associations between VSLY quintiles and covariates.

| Variable | Group | VSLY |  |  |  |  | *p* value |
| --- | --- | --- | --- | --- | --- | --- | --- |
|  |  | Q1<br>(563) | Q2<br>(556) | Q3<br>(546) | Q4<br>(544) | Q5<br>(552) |  |
| Frailty | Non-frail | 197<br>[35.0%] | 214<br>[38.5%] | 224<br>[41.0%] | 247<br>[45.4%] | 290<br>[52.5%] | <0.001* |
|  | Pre-frail and Frail | 366<br>[65.0%] | 342<br>[61.5%] | 322<br>[59.0%] | 297<br>[54.6%] | 262<br>[47.5%] |  |
| Gender | Male | 241<br>[42.8%] | 250<br>[45.0%] | 242<br>[44.3%] | 274<br>[50.4%] | 60.5% | <0.001* |
|  | Female | 322<br>[57.2%] | 306<br>[55.0%] | 304<br>[55.7%] | 270<br>[49.6%] | 39.5% |  |
| Marital Status | Married | 396<br>[70.3%] | 411<br>[73.9%] | 398<br>[72.9%] | 414<br>[76.1%] | 458<br>[83.0%] | <0.001* |
|  | Single | 167<br>[29.7%] | 145<br>[26.1%] | 148<br>[27.1%] | 130<br>[23.9%] | 94<br>[17.0%] |  |
| Education | No education | 485<br>[86.1%] | 473<br>[85.1%] | 464<br>[85.0%] | 470<br>[86.4%] | 442<br>[80.1%] | 0.025* |
|  | Any education | 78<br>[13.9%] | 83<br>[14.9%] | 82<br>[15.0%] | 74<br>[13.6%] | 110<br>[19.9%] |  |
| Wealth quintiles<br>(Q5 is the highest) | Q1 | 126<br>[22.4%] | 102<br>[18.3%] | 109<br>[20.0%] | 106<br>[19.5%] | 99<br>[17.9%] | 0.855<sup>#</sup> |
|  | Q2 | 116<br>[20.6%] | 113<br>[20.3%] | 82<br>[15.0%] | 122<br>[22.4%] | 115<br>[20.8%] |  |
|  | Q3 | 103<br>[18.3%] | 96<br>[17.3%] | 123<br>[22.5%] | 114<br>[21.0%] | 115<br>[20.8%] |  |
|  | Q4 | 105<br>[18.7%] | 123<br>[22.1%] | 120<br>[22.0%] | 103<br>[18.9%] | 118<br>[21.4%] |  |
|  | Q5 | 113<br>[20.1%] | 122<br>[21.9%] | 112<br>[20.5%] | 99<br>[18.2%] | 105<br>[19.0%] |  |
| Multimorbidity | No | 75.3% | 72.8% | 79.3% | 78.3% | 82.1% | 0.003* |
|  | Yes | 27.2% | 20.7% | 21.7% | 17.9% | 17.9% |  |
| Age | Correlation coefficient | -0.054 (-0.092,-0.016) |  |  |  |  | 0.004<sup>^</sup> |
| Quality of life (score) | Correlation coefficient | -0.036 (-0.074,-0.002) |  |  |  |  | 0.058<sup>^</sup> |
| Disability (score) | Correlation coefficient | -0.069 (-0.107_-0.030) |  |  |  |  | <0.001<sup>^</sup> |

\* Pearson Chi-Square
^ Spearman's correlation

Multicollinearity between variables was excluded. Sequential binary logistic regression models showed a significantly greater odds having a high VSLY in those who were non-frail compared to frail or pre-frail. This became non-significant for the lower VSLY quintiles (Table 5). The inverse linear relationship between frailty and VSLY wasn’t substantially affected by adding socio-demographic or economic variables, or quality of life, or multimorbidity, or disability to the model (Table 5). Forest plots showing the relationship between frailty and VSLY for each set of quintiles comparisons in each sequential model are shown in Appendix Fig. 2, Supplementary Information 3. Frailty ORs from the fully adjusted sequential model (Model 5) in each comparison of VSLY quintiles are shown in Figure 3.

**Figure 3.**
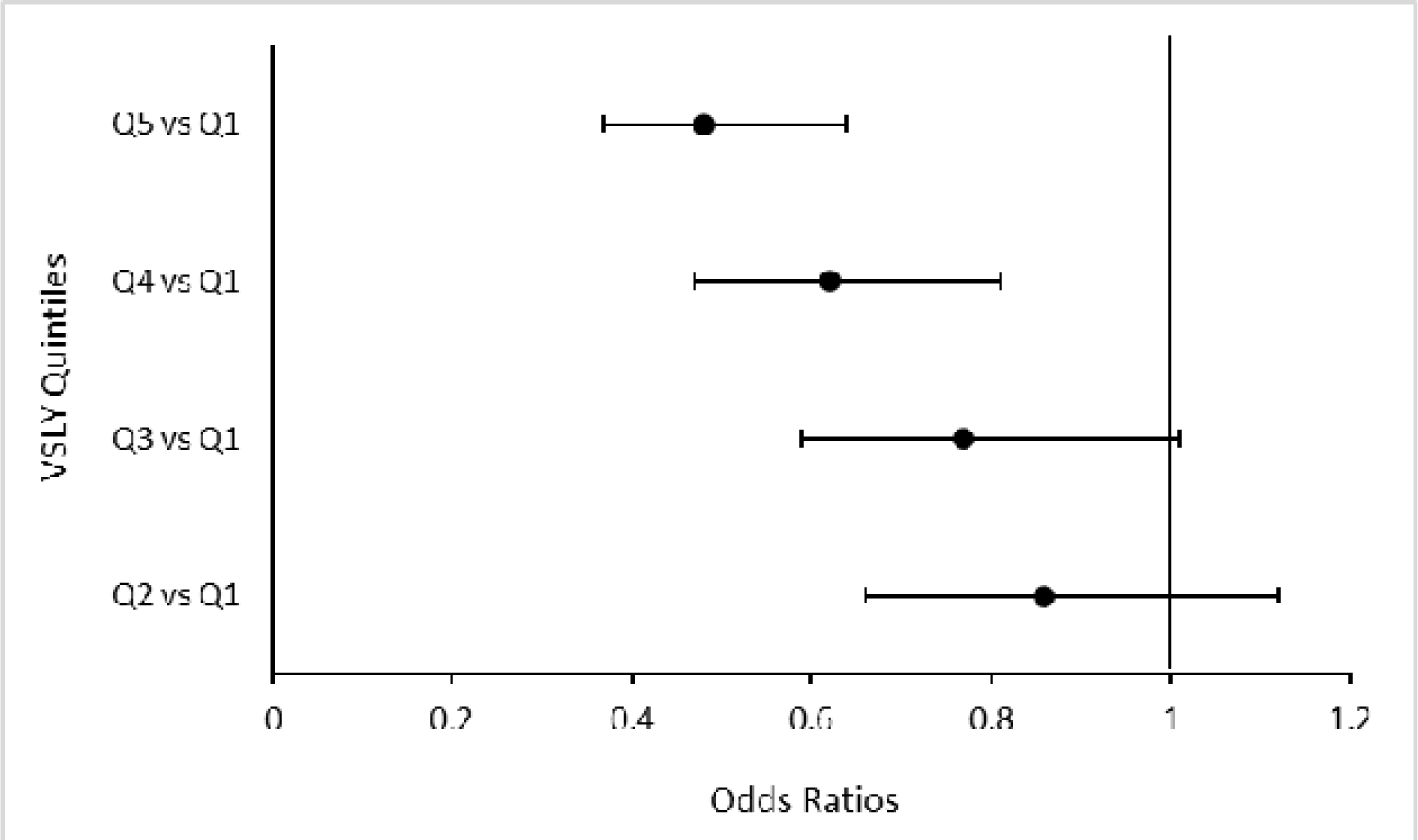
Forest plot representing the Odd Ratios for frailty in each comparison of VSLY quintiles, controlled for all covariates.

**Table 5.**
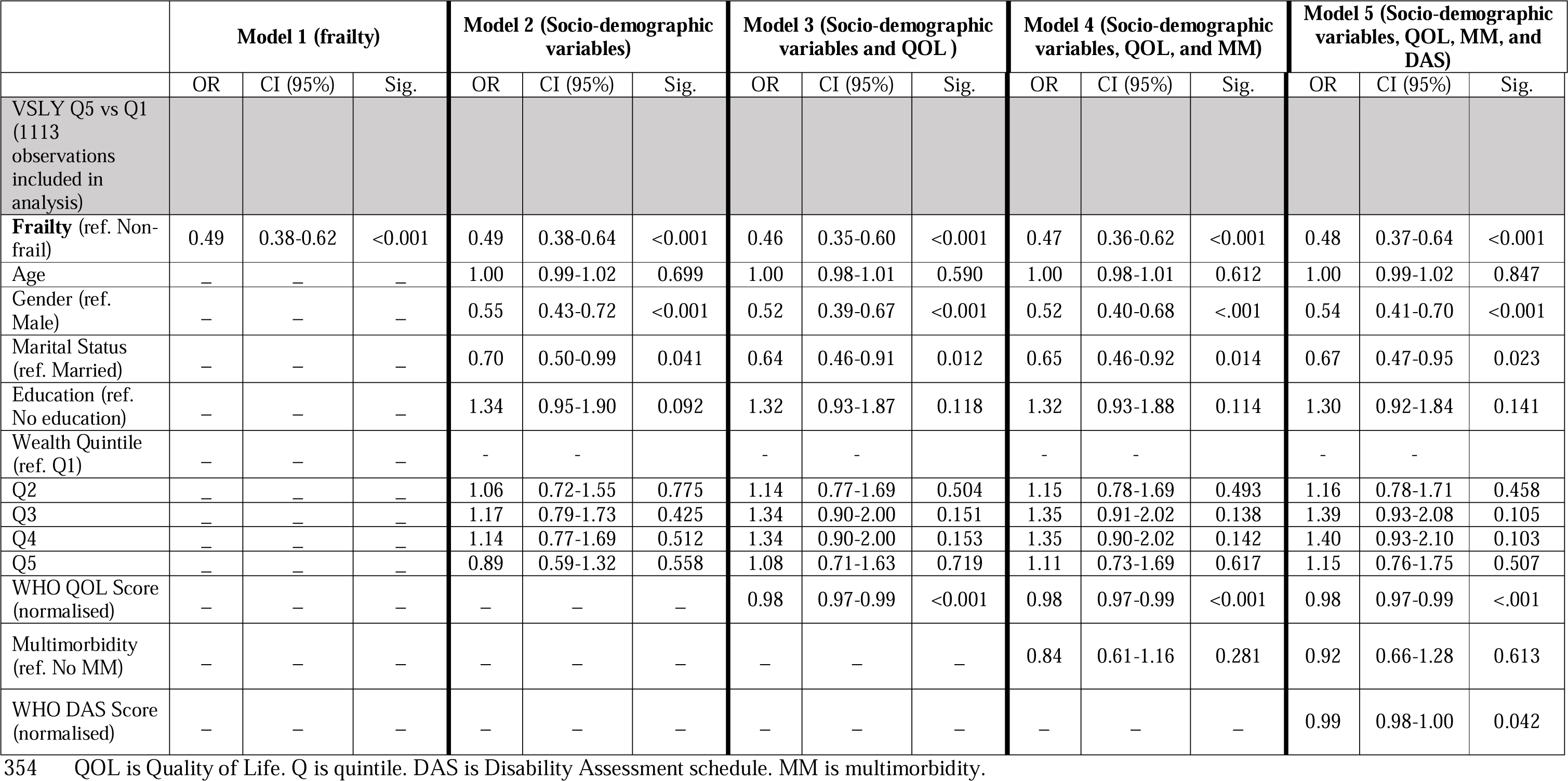
Sequential model adjustments for comparing the highest VSLY quintile (Q5) with the first one (Q1), investigating the association with frailty, quality of life, and multimorbidity after controlling for other confounding variables. All quartile comparisons of VSLY are shown in **Appendix Table 2, Supplementary Information 3.**

In addition to frailty, being female or single were significantly associated with a lower VSLY whereas being older, educated, or wealthy had no statistically significant association. These associations were not substantially modified by adding quality of life, multimorbidity, disability, or all together to the model. Higher quality of life was significantly associated with a lower VSLY, greater disability was associated with lower VSLY, whereas there was no significant association between VSLY and multimorbidity (see Table 5 and Appendix Table 2, Supplementary Information 3).

The results for the multinomial regression done for the sensitivity analyses were not substantially different to the binary logistic regression models (See Appendix Table 3 and Appendix Fig. 3, Supplementary Information 3).

## 4. Discussion

We found a strong and significant association between frailty and VSLY in this population in Burkina Faso with people who are frail assigning lower values to prevent a mortality risk than those who are non-frail (robust). This association persisted after adjustment for other variables found in other studies likely to impact upon the relationship, such as quality of life, multimorbidity, and age [17–19].

Given the global and widespread use of VSLY approaches to indicate benefits and calls to use this or similar approaches to inform investment cases for health, it is important to understand how conditions which affect health of populations in whom VSLY might be calculated impact upon it [5, 33]. Our findings have relevance to the interpretation of VSLY values in older people in low-income settings and will become increasingly important to recognise as ageing populations increase in these settings. As far as we are aware, this is the first study to assess implications of frailty on VSLY in any setting, hence our results have applicability for many countries where VSLY is being considered for use in an older population.

When frailty was included in the model, the effect of age, wealth, or education on VSLY – which were strongly associated with VSLY on bivariate analyses, became non-significant. Age and wealth have been shown to be associated with VSLY in other studies but when controlling for other variables in these studies these relationships were attenuated [9], as they did in our study. In fact, that frailty has such a strong association with VSLY—over and above that of other variables traditionally thought of as being associated with VSLY—suggests that it may be necessary to assess frailty status when estimating VSLY. This is particularly important given that frailty is a highly dynamic state, thus VSLY captured in frail individuals may change if that person changes frailty status [34].

Although not our main variables of interest, we saw associations between VSLY and other covariates that are worth discussing. Similar to our study, previous studies have shown a relationship between gender and VSLY that persists despite controlling for covariates. Women consistently value their lives lower than do men [35]. Our finding that being married or cohabitating is associated with odds of a higher VSLY is novel, but unsurprising if it is hypothesised that having family responsibilities may increase the willingness to pay to avoid risk. Another study showed that married respondents have significantly higher WTP to take care of a child/other adult than to protect themselves [18]. The relationship between greater disability and lower willingness to pay was small, but significant. Again, this finding is not surprising, given the influence of disability on people’s perceived value [36–38]. However, our finding that increasing quality of life is associated, both in bivariate and fully adjusted analyses, with lower VSLY is puzzling. We hypothesise that this might be due to older people with a higher quality of life also being comfortable with their status quo, but this hypothesis requires further investigation in qualitative studies. That multimorbidity was associated with lower VSLY in bivariate analyses but not the fully adjusted model suggests that the impact of MM on VSLY may be mostly mediated through frailty and somewhat through disability.

Our study has several limitations. Given the cross-sectional nature of the study, we can only determine associations and lack the longitudinal data needed for determining causation. We grouped frail and pre-frail individuals into one category, losing the nuance of showing effects by three categories. However, this has been done in to aid in the interpretation of results. We have also shown (personal communication) that these categories behave in a similar way when considering future outcomes of this population, so grouping them together is reasonable. The number of conditions that we used to derive multimorbidity was limited by questions asked in the survey, but they have been used in previous publications to classify multimorbidity [15]. Limited sample sizes for the individual VSLY methods necessitated combining the data using quintiles; however, reassuringly, we saw similar relationships between frailty and VSLY when the analysis was done by individual VSLY methodologies. If the inverse U shape of the association with age found in other studies, was found in our study, our adjustment for age may have introduced some errors. However, in our sample, we found a linear association between VSLY and age. We used the method of Patenaude [6] to estimate VSLY, as it has been found suitable for use in other similar settings [39]. However, we acknowledge that there are other methods that could have been applied. Finally, our study lacks the contextual grounding which would be afforded by doing mixed methods analysis and qualitative exploration to allow a deeper understanding of the context. Future studies should explore what is important to people in the Nouna region of Burkina Faso in terms of what they value in life, and how they relate their values to money.

Our analysis also has several strengths. There have not been many former attempts to study associations with VSLY in LMICs, and none that we are aware of globally which have ascertained associations with frailty in an ageing population. We used multiple ways of assessing VSLY in a fairly large sample, and included a broad range of covariates into the analysis. We used a standard way of evaluating frailty, including use of objective measures of physical performance; standard QOL and disability tools, facilitating replication in other settings.

In conclusion, ageing is a neglected phenomenon in many LMICs, where research has tended to focus on diseases of poverty and infectious diseases more common in younger people. This is especially the case in low income countries (LICs), which are typically thought of as being less far along the epidemiological transition [7]. However, our recent work has shown that populations in LICs suffer a high burden of multimorbidity, which is associated with frailty [40, 41]. Whilst our findings suggest that people who are frail place a lower value on their lives, and given that frailty status is dynamic, we recommend that frailty status is taken into account when calculating VSLY to avoid older people being “assigned” lower values than they should be.

## Declarations

### Availability of data and material

Data may be obtained from a third party and are not publicly available. Data are not publicly available as consent was not given by participants for this to take place. This is in part because entire age cohorts of some villages are included in the data set, potentially allowing for deductive disclosure with sufficient local information. For this reason, anonymised data are available from CHAS study data controllers only following signature of a data use agreement restricting onward transmission. Anyone wishing to replicate the analyses presented or conduct further collaborative analyses using CHAS (which are welcomed and considered based on a letter of intent), should contact Prof Justine Davies in the first instance.

### Ethics approval

Ethical approval was obtained from Ethics Commission I of the medical faculty Heidelberg (S-120/2018), the Burkina Faso Comité d’Ethique pour la Recherche en Santé (CERS) in Ouagadougou (2018-4-045) and the Institutional Ethics Committee (CIE) of the CRSN (2018-04).

### Consent to participate

Oral consent was obtained from all village elders. Written informed consent was obtained from each participant and a literate witness assisted in cases of illiteracy. All participants with test results indicating requirement for healthcare were contacted and provided with referral to the appropriate level of care.

### Consent for publication

Not applicable Code availability Not applicable

### Author contributions

JD, MW, ST, JMG, GH, and PG conceived and designed the overall CSRN CHAS study. GH co-ordinated baseline data collection and preparation. LF, DGP, FB, JID, and MDW designed the current study. LF conducted the analysis, wrote, and revised the manuscript. JD, DGP, and MW supervised the analysis, write up and development of the manuscript. All authors substantively reviewed manuscripts, inputted into revisions and approved the final manuscript.

## Data Availability

All data produced in the present study are available upon reasonable request to the authors.

## Acknowledgements

Not applicable

## List of abbreviations

VSLY: Value of Statistical Life Years
LMICs: Low- and Middle-Income Countries
MM: Multimorbidity
QOL: Quality Of Life
DAS: Disability Assessment Schedule

