## Supplementary Materials for "The association between the value of a statistical life and frailty in Burkina Faso"

### Appendix 1 - Details of assessment of VSLY in Nouna

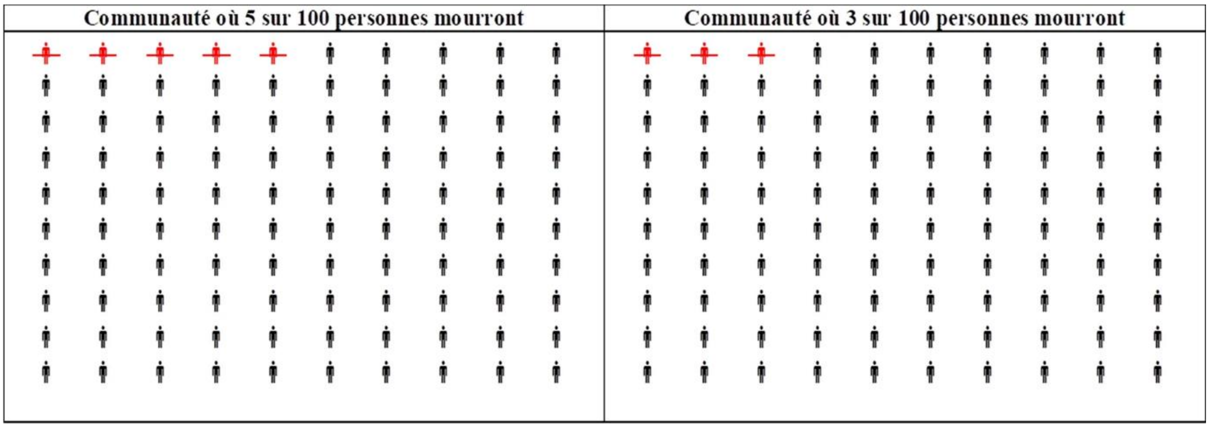


**Appendix Fig. 1** Graphical representation of health risks, from 'Value of statistical life year in extreme poverty: a randomized experiment of measurement methods in rural Burkina Faso', Popul Health Metrics 19, 45 (2021) [1]

Given the very low literacy levels in Nouna, risk changes were chosen to allow meaningful communication; a feasible design for very low-income contexts that was first developed by Patenaude et al. (2019) [2]. During data collection, interviewers described two identical communities (with 100 people in each) which only differed in terms of the probability that each of its members may suffer a sudden and painless death in each year (Appendix Fig. 1). Respondents were told to imagine being one of these 100 people in one of these communities (lower risk community for WTA; larger-risk community for WTP), without knowing whether they will be one of those who live or die. They were then asked to specify the annual price they would pay to move to the lower-risk community (WTP), or the payment they require to move to the larger risk community (WTA).

#### Appendix 2 - Derivation of frailty scores

Weight loss was defined as self-reported loss of >4 kg over the last 12 months, low activity levels were defined as the highest quintile of self-reported hours of sitting per week for each sex. Self-reported exhaustion was taken as a positive response was to either statement, as above, from the CES-D instrument applying for at least 3–4 days per week. Low grip strength was the lowest quintile of BMI-adjusted grip for each sex and low walk speed was the lowest quintile of height-adjusted walk speed. Each domain (low grip strength, low walk speed, or self-reported weight loss, exhaustion, or low activity levels) scored 1 point giving a final total between 0 and 5 points. Participants with missing data on one or more of the physical assessment measures were categorised as 'Frail' based on previous work in South Africa.

**Appendix Table* 1** Equations and cut-off values used to derive Fried frailty score

| | **Men** | **Women** |
|---|---|---|
| Mean grip strength (kg) | 44.9 | 31.1 |
| Equation for adjusted grip strength | = maximum grip - ((BMI - 21.6) * 1.17). | = maximum grip - ((BMI – 22.4) * 0.44). |
| Low BMI-adjusted grip strength | <37.1kg | <25.0kg |
| Mean walk speed (m/s) | 1.045 | 0.888 |
| Equation for adjusted walk speed | = walk speed - ((height in cm-172) * 0.00579) | = walk speed - ((height in cm - 162) * 0.00849) |
| Low height adjusted walk speed | <0.821m/s | <0.720m/s |
| Self-reported weight loss | >4kg in last year | >4kg in last year |
| Self-reported exhaustion (2 questions) | Response of '3 to 4 days a week' or more often to either question | Response of '3 to 4 days a week' or more often to either question |
| Self-reported low activity levels (denoted by hours sitting per week) | >=34 hours | >=36 hours |

BMI, Body Mass Index.

* *Table taken with permission from: 'Epidemiology of multimorbidity in conditions of extreme poverty: a population-based study of older adults in rural Burkina Faso', BMJ Global Health 2020; 5:e002096 [1].*

#### Appendix 3 – Details on sequential models of the relationship between frailty and VSLY

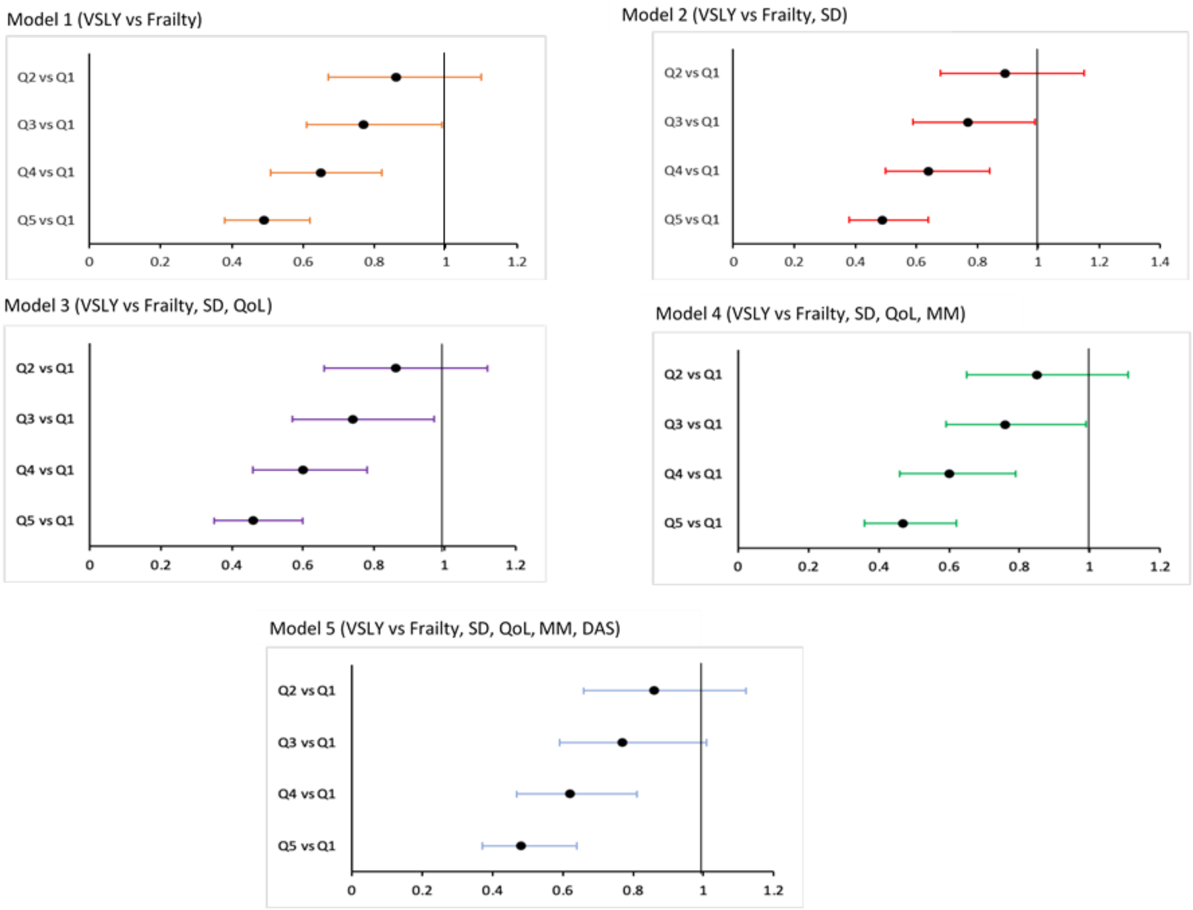


**Appendix Fig. 2** Forest plots of relationships between VSLY and frailty for each set of quintiles comparisons in each sequential binary logistic model. DAS, Disability Assessment Schedule; MM, MultiMorbidity; QoL, Quality of Life; SD, Sociodemographic variables; VSLY, Value of a Statistical Life Year

**Appendix Table 2** Results of binary logistic regression comparing each VSLY quintile with the first one in sequential models

| **Compared Quintiles** | **Model 1 (frailty)** | | | **Model 2 (Socio-demographic variables)** | | | **Model 3 (Socio-demographic variables aand QoL )** | | | **Model 4 (Socio-demographic variables, QoL, and Multimorbidity)** | | | **Model 5 (Socio-demographic variables, QoL, MM, and DAS)** | | |
|---|---|---|---|---|---|---|---|---|---|---|---|---|---|---|---|
| | OR | CI (95%) | Sig. | OR | CI (95%) | Sig. | OR | CI (95%) | Sig. | OR | CI (95%) | Sig. | OR | CI (95%) | Sig. |
| Q5 vs Q1 (1113 observations included in analysis) | | | | | | | | | | | | | | | |
| **Frailty** (ref. Non-frail) | 0.49 | 0.38-0.62 | <0.001 | 0.49 | 0.38-0.64 | <0.001 | 0.46 | 0.35-0.60 | <0.001 | 0.47 | 0.36-0.62 | <0.001 | 0.48 | 0.37-0.64 | <0.001 |
| Age | _ | _ | _ | 1.00 | 0.99-1.02 | 0.699 | 1.00 | 0.98-1.01 | 0.590 | 1.00 | 0.98-1.01 | 0.612 | 1.00 | 0.99-1.02 | 0.847 |
| Gender (ref. Male) | _ | _ | _ | 0.55 | 0.43-0.72 | <0.001 | 0.52 | 0.39-0.67 | <0.001 | 0.52 | 0.40-0.68 | <.001 | 0.54 | 0.41-0.70 | <0.001 |
| Marital Status (ref. Married) | _ | _ | _ | 0.70 | 0.50-0.99 | 0.041 | 0.64 | 0.46-0.91 | 0.012 | 0.65 | 0.46-0.92 | 0.014 | 0.67 | 0.47-0.95 | 0.023 |
| Education (ref. No education) | _ | _ | _ | 1.34 | 0.95-1.90 | 0.092 | 1.32 | 0.93-1.87 | 0.118 | 1.32 | 0.93-1.88 | 0.114 | 1.30 | 0.92-1.84 | 0.141 |
| Wealth Quintile (ref. Q1) | _ | _ | _ | - | - | | - | - | | - | - | | - | - | |
| Q2 | _ | _ | _ | 1.06 | 0.72-1.55 | 0.775 | 1.14 | 0.77-1.69 | 0.504 | 1.15 | 0.78-1.69 | 0.493 | 1.16 | 0.78-1.71 | 0.458 |
| Q3 | _ | _ | _ | 1.17 | 0.79-1.73 | 0.425 | 1.34 | 0.90-2.00 | 0.151 | 1.35 | 0.91-2.02 | 0.138 | 1.39 | 0.93-2.08 | 0.105 |
| Q4 | _ | _ | _ | 1.14 | 0.77-1.69 | 0.512 | 1.34 | 0.90-2.00 | 0.153 | 1.35 | 0.90-2.02 | 0.142 | 1.40 | 0.93-2.10 | 0.103 |
| Q5 | _ | _ | _ | 0.89 | 0.59-1.32 | 0.558 | 1.08 | 0.71-1.63 | 0.719 | 1.11 | 0.73-1.69 | 0.617 | 1.15 | 0.76-1.75 | 0.507 |
| WHO QOL Score (normalised) | _ | _ | _ | _ | _ | _ | 0.98 | 0.97-0.99 | <0.001 | 0.98 | 0.97-0.99 | <0.001 | 0.98 | 0.97-0.99 | <.001 |
| Multimorbidity (ref. No MM) | _ | _ | _ | _ | _ | _ | _ | _ | _ | 0.84 | 0.61-1.16 | 0.281 | 0.92 | 0.66-1.28 | 0.613 |

| | | | | | | | | | | | | | | | |
|---|---|---|---|---|---|---|---|---|---|---|---|---|---|---|---|
| WHO DAS Score (normalised) | – | – | – | – | – | – | – | – | – | – | – | – | 0.99 | 0.98-1.00 | 0.042 |
| Q4 vs Q1 (1106 observations included in analysis) | | | | | | | | | | | | | | | |
| **Frailty** (ref. Robust) | 0.65 | 0.51-0.82 | <0.001 | 0.64 | 0.50-0.84 | 0.001 | 0.600 | 0.46-0.78 | <0.001 | 0.61 | 0.46-0.79 | <0.001 | 0.62 | 0.47-0.81 | <.001 |
| Age | – | – | – | 1.00 | 0.99-1.01 | 0.549 | 0.997 | 0.98-1.01 | 0.689 | 1.00 | 0.98-1.01 | 0.733 | 1.00 | 0.99-1.01 | 0.861 |
| Gender (ref. Male) | – | – | – | 0.76 | 0.59-0.99 | 0.038 | 0.729 | 0.56-0.94 | 0.017 | 0.73 | 0.56-0.95 | .019 | 0.74 | 0.57-0.95 | 0.021 |
| Marital Status (ref. Married) | – | – | – | 0.87 | 0.64-1.19 | 0.394 | 0.847 | 0.62-1.16 | 0.306 | 0.85 | 0.62-1.17 | 0.326 | 0.87 | 0.63-1.19 | 0.385 |
| Education (ref. No education) | – | – | – | 0.93 | 0.65-1.34 | 0.695 | 0.927 | 0.64-1.34 | 0.687 | 0.93 | 0.65-1.35 | 0.719 | 0.92 | 0.63-1.33 | 0.643 |
| Wealth Q (ref. Q1) | – | – | – | - | - | | | | | | | | | | |
| Q2 | – | – | – | 1.18 | 0.82-1.71 | 0.370 | 1.27 | 0.875-1.84 | 0.207 | 1.27 | 0.88-1.85 | 0.204 | 1.28 | 0.88-1.86 | 0.194 |
| Q3 | – | – | – | 1.29 | 0.88-1.88 | 0.189 | 1.41 | 0.962-2.08 | 0.078 | 1.42 | 0.97-2.09 | 0.075 | 1.43 | 0.97-2.10 | 0.072 |
| Q4 | – | – | – | 1.10 | 0.75-1.60 | 0.639 | 1.25 | 0.85-1.86 | 0.250 | 1.26 | 0.85-1.86 | 0.246 | 1.28 | 0.87-1.90 | 0.213 |
| Q5 | – | – | – | 0.97 | 0.66-1.43 | 0.889 | 1.14 | 0.76-1.70 | 0.515 | 1.16 | 0.78-1.74 | 0.459 | 1.19 | 0.79-1.79 | 0.396 |
| WHO QOL Score (normalised) | – | – | – | – | – | – | 0.98 | 0.97-0.99 | <0.001 | 0.98 | 0.97-0.99 | <0.001 | 0.98 | 0.97-0.99 | <0.001 |
| Multimorbidity (ref. No MM) | – | – | – | – | – | – | – | – | – | 0.89 | 0.66-1.21 | 0.466 | 0.96 | 0.70-1.31 | 0.784 |
| WHO DAS Score (normalised) | – | – | – | – | – | – | – | – | – | – | – | – | 0.99 | 0.98-1.00 | 0.145 |
| Q3 vs Q1 (1108 observations included in analysis) | | | | | | | | | | | | | | | |

| | | | | | | | | | | | | | | | |
|---|---|---|---|---|---|---|---|---|---|---|---|---|---|---|---|
| **Frailty** (ref. Non-frail) | 0.77 | 0.61-0.99 | 0.039 | 0.77 | 0.59-0.99 | 0.045 | 0.74 | 0.57-0.97 | 0.028 | 0.76 | 0.59-0.99 | 0.046 | 0.77 | 0.59-1.01 | 0.056 |
| Age | _ | _ | _ | 1.00 | 0.99-1.01 | 0.835 | 1.00 | 0.98-1.01 | 0.661 | 1.00 | 0.99-1.01 | 0.794 | 1.00 | 0.99-1.01 | 0.979 |
| Gender (ref. Male) | _ | _ | _ | 0.98 | 0.76-1.27 | 0.907 | 0.96 | 0.74-1.24 | 0.750 | 0.97 | 0.75-1.26 | 0.824 | 0.98 | 0.76-1.27 | 0.886 |
| Marital Status (ref. Married) | _ | _ | _ | 0.94 | 0.69-1.28 | 0.709 | 0.91 | 0.67-1.24 | 0.569 | 0.92 | 0.67-1.25 | 0.601 | 0.93 | 0.68-1.27 | 0.661 |
| Education (ref. No education) | _ | _ | _ | 1.07 | 0.750-1.52 | 0.713 | 1.07 | 0.750-1.53 | 0.713 | 1.08 | 0.75-1.54 | 0.683 | 1.06 | 0.74-1.52 | 0.741 |
| Wealth Q (ref. Q1) | _ | _ | _ | - | - | | | | | | | | | | |
| Q2 | _ | _ | _ | 0.76 | 0.52-1.13 | 0.178 | 0.81 | 0.54-1.19 | 0.283 | 0.82 | 0.55-1.22 | 0.202 | 0.83 | 0.56-1.22 | 0.356 |
| Q3 | _ | _ | _ | 1.32 | 0.91-1.92 | 0.146 | 1.43 | 0.97-2.09 | 0.067 | 1.46 | 1.00-2.14 | 0.133 | 1.48 | 1.01-2.17 | 0.045 |
| Q4 | _ | _ | _ | 1.24 | 0.85-1.80 | 0.267 | 1.34 | 0.93-2.01 | 0.111 | 1.41 | 0.95-2.07 | 0.215 | 1.43 | 0.97-2.12 | 0.071 |
| Q5 | _ | _ | _ | 1.07 | 0.73-1.57 | 0.726 | 1.21 | 0.81-1.79 | 0.347 | 1.27 | 0.85-1.89 | 0.559 | 1.30 | 0.87-1.95 | 0.194 |
| WHO QOLScore (normalised) | _ | _ | _ | _ | _ | _ | 0.99 | 0.98-1.00 | 0.008 | 0.99 | 0.98-1.00 | 0.004 | 0.98 | 0.97-0.99 | 0.002 |
| Multimorbidity (ref. No MM) | _ | _ | _ | _ | _ | _ | _ | _ | _ | 0.77 | 0.57-1.04 | 0.084 | 0.80 | 0.59-1.09 | 0.164 |
| WHO DAS Score (normalised) | _ | _ | _ | _ | _ | _ | _ | _ | _ | _ | _ | _ | 1.00 | 0.99-1.00 | 0.340 |
| Q2 vs Q1 (1118 observations included in analysis) | | | | | | | | | | | | | | | |
| **Frailty** (ref. Non-frail) | 0.86 | 0.67-1.10 | 0.225 | 0.89 | 0.68-1.15 | 0.367 | 0.86 | 0.66-1.12 | 0.276 | 0.85 | 0.65-1.11 | 0.229 | 0.86 | 0.66-1.12 | 0.263 |
| Age | _ | _ | _ | 1.00 | 0.99-1.01 | 0.716 | 1.00 | 0.99-1.01 | 0.963 | 1.00 | 0.99-1.01 | 0.941 | 1.00 | 0.99-1.01 | 0.759 |
| Gender (ref. Male) | _ | _ | _ | 0.97 | 0.75-1.25 | 0.826 | 0.96 | 0.74-1.24 | 0.744 | 0.95 | 0.74-1.23 | 0.714 | 0.96 | 0.74-1.24 | 0.769 |

| | | | | | | | | | | | | | | | |
|---|---|---|---|---|---|---|---|---|---|---|---|---|---|---|---|
| Marital Status (ref. Married) | – | – | – | 0.88 | 0.65-1.20 | 0.416 | 0.87 | 0.64-1.18 | 0.359 | 0.86 | 0.63-1.17 | 0.335 | 0.87 | 0.64-1.19 | 0.395 |
| Education (ref. No education) | – | – | – | 1.03 | 0.72-1.47 | 0.880 | 1.02 | 0.72-1.46 | 0.898 | 1.03 | 0.72-1.47 | 0.880 | 1.02 | 0.71-1.46 | 0.924 |
| Wealth Q (ref. Q1) | - | - | - | - | - | - | - | - | - | - | - | - | - | - | - |
| Q2 | – | – | – | 1.15 | 0.79-1.67 | 0.463 | 1.18 | 0.81-1.73 | 0.377 | 1.18 | 0.81-1.72 | 0.394 | 1.20 | 0.82-1.75 | 0.341 |
| Q3 | – | – | – | 1.11 | 0.75-1.64 | 0.597 | 1.16 | 0.78-1.72 | 0.462 | 1.14 | 0.77-1.70 | 0.511 | 1.17 | 0.78-1.74 | 0.443 |
| Q4 | – | – | – | 1.37 | 0.94-2.00 | 0.101 | 1.44 | 0.98-2.12 | 0.062 | 1.43 | 0.97-2.10 | 0.071 | 1.47 | 1.00-2.16 | 0.052 |
| Q5 | – | – | – | 1.27 | 0.86-1.85 | 0.226 | 1.35 | 0.91-2.00 | 0.134 | 1.30 | 0.88-1.94 | 0.191 | 1.35 | 0.90-2.03 | 0.140 |
| WHO QOL Score (normalised) | – | – | – | – | – | – | 0.99 | 0.98-1.00 | 0.166 | 0.99 | 0.98-1.00 | 0.211 | 0.99 | 0.98-1.00 | 0.128 |
| Multimorbidity (ref. No MM) | – | – | – | – | – | – | – | – | – | 1.16 | 0.88-1.55 | 0.296 | 1.22 | 0.91-1.65 | 0.187 |
| WHO DAS Score (normalised) | – | – | – | – | – | – | – | – | – | – | – | – | 0.99 | 0.99-1.00 | 0.295 |

DAS, Disability Assessment schedule; MM, multimorbidity; Q is quintile; QOL, Quality of Life

**Appendix Table 3** Results of fully adjusted multinomial model

| **Compared Quintile** | **OR** | **CI (95%)** | **Sig.** |
|---|---|---|---|
| Q5 | | | |
| **Frailty** (ref. Non-frail) | 0.51 | 0.39-0.66 | <0.001 |
| Age | 1.00 | 0.99-1.01 | 0.999 |
| Gender (ref. Male) | 0.55 | 0.42-0.72 | <0.001 |
| Marital Status (ref. Married) | 0.72 | 0.52-1.01 | 0.058 |
| Education (ref. No education) | 1.28 | 0.91-1.81 | 0.150 |
| Q1 | 0.85 | 0.56-1.28 | 0.425 |
| Q2 | 0.99 | 0.67-1.46 | 0.952 |
| Q3 | 1.16 | 0.78-1.72 | 0.468 |
| Q4 | 1.18 | 0.80-1.74 | 0.400 |
| Wealth Q (ref. Q5) | - | - | - |
| WHO QOL Score (normalised) | 0.98 | 0.97-0.99 | <0.001 |
| Multimorbidity (ref. No MM) | 0.91 | 0.66-1.26 | 0.571 |
| WHO DAS Score (normalised) | 0.99 | 0.98-1.00 | 0.017 |
| Q4 | | | |
| **Frailty** (ref. Robust) | 0.63 | 0.49-0.83 | <0.001 |
| Age | 1.00 | 0.99-1.01 | 0.876 |
| Gender (ref. Male) | 0.74 | 0.57-0.96 | 0.026 |
| Marital Status (ref. Married) | 0.87 | 0.63-1.19 | 0.374 |
| Education (ref. No education) | 0.91 | 0.63-1.31 | 0.609 |
| Q1 | 0.83 | 0.55-1.24 | 0.355 |
| Q2 | 1.04 | 0.71-1.54 | 0.829 |
| Q3 | 1.14 | 0.77-1.69 | 0.503 |
| Q4 | 1.03 | 0.70-1.53 | 0.863 |
| Wealth Q (ref. Q5) | - | - | - |
| WHO QOL Score (normalised) | 0.98 | 0.97-0.99 | <0.001 |
| Multimorbidity (ref. No MM) | 0.97 | 0.71-1.32 | 0.833 |
| WHO DAS Score (normalised) | 0.99 | 0.98-1.00 | 0.160 |
| Q3 | | | |
| **Frailty** (ref. Non-frail) | 0.78 | 0.60-1.01 | 0.058 |

| | | | |
|---|---|---|---|
| Age | 1.00 | 0.99-1.01 | 0.953 |
| Gender (ref. Male) | 0.96 | 0.74-1.25 | 0.785 |
| Marital Status (ref. Married) | 0.94 | 0.69-1.29 | 0.717 |
| Education (ref. No education) | 1.05 | 0.73-1.49 | 0.795 |
| Q1 | 0.75 | 0.50-1.11 | 0.154 |
| Q2 | 0.63 | 0.42-0.95 | 0.026 |
| Q3 | 1.10 | 0.75-1.62 | 0.610 |
| Q4 | 1.08 | 0.74-1.58 | 0.689 |
| Wealth Q (ref. Q5) | - | - | - |
| WHO QOLScore (normalised) | 0.98 | 0.97-0.99 | 0.002 |
| Multimorbidity (ref. No MM) | 0.80 | 0.59-1.09 | 0.162 |
| WHO DAS Score (normalised) | 1.00 | 0.99-1.00 | 0.355 |
| Q2 | | | |
| **Frailty** (ref. Non-frail) | 0.86 | 0.66-1.12 | 0.262 |
| Age | 1.00 | 0.99-1.02 | 0.778 |
| Gender (ref. Male) | 0.95 | 0.73-1.23 | 0.709 |
| Marital Status (ref. Married) | 0.87 | 0.64-1.18 | 0.370 |
| Education (ref. No education) | 1.02 | 0.72-1.45 | 0.912 |
| Q1 | 0.74 | 0.50-1.10 | 0.132 |
| Q2 | 0.89 | 0.61-1.31 | 0.561 |
| Q3 | 0.86 | 0.58-1.27 | 0.444 |
| Q4 | 1.08 | 0.75-1.57 | 0.673 |
| Wealth Q (ref. Q5) | - | - | - |
| WHO QOLScore (normalised) | 0.99 | 0.98-1.00 | 0.165 |
| Multimorbidity (ref. No MM) | 1.20 | 0.89-1.61 | 0.232 |
| WHO DAS Score (normalised) | 1.00 | 0.99-1.00 | 0.385 |

DAS, Disability Assessment schedule; MM, multimorbidity; Q is quintile; QOL, Quality of Life

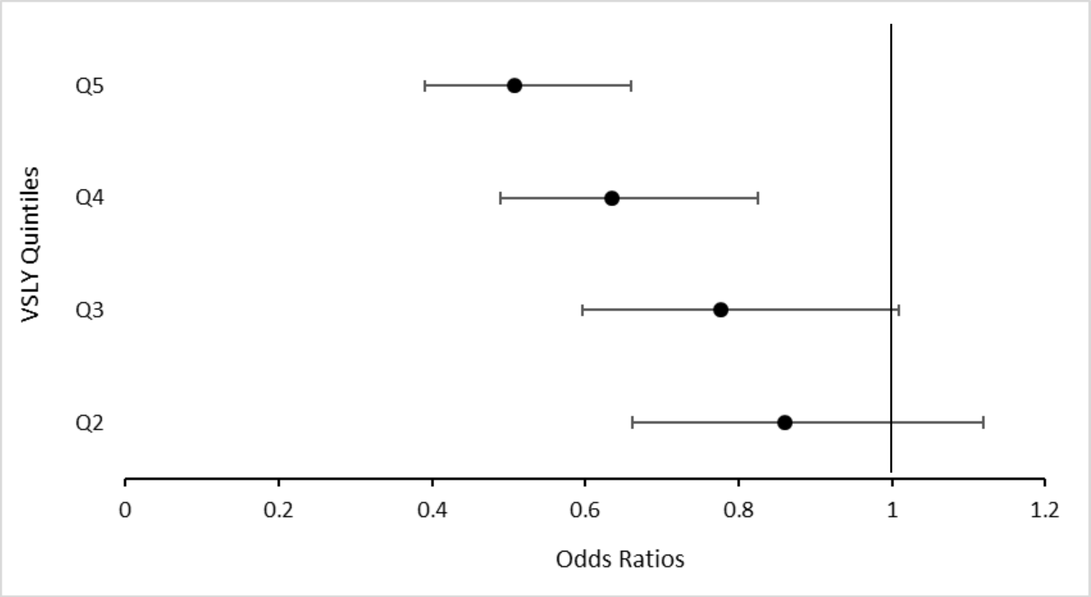


**Appendix Fig. 3** Forest plots of relationships between VSLY and frailty for each set of quintiles comparisons in multinomial regression model (sensitivity analyses)
